# Characterization of dynamic postural control during weight load shifting with and without support surface reduction

**DOI:** 10.64898/2026.04.30.26352157

**Authors:** Esteban Osella, Ricardo Alfredo Rettore, Paola Catalfamo, J. A. Biurrun Manresa, Yanina Verónica Atum

## Abstract

**Purpose:** to characterize the dynamic postural control during weight load shifting with and without support surface reduction with temporal metrics commonly used in linear control systems identification.

**Methods:** From the COP coordinates temporal, global and structural parameters were calculated. Reliability of derived parameters were determined using Bland-Altman analysis.

**Results:** For the observed population, temporal variables tend to decrease when the complexity of the task is increased with the reduction in the support surface and the non dominance.

**Conclusion:** Delay and rise times were significantly shorter for the non-dominant limb in the anteroposterior direction when volunteers performed the same task with different limbs. In the mediolateral direction, delay and rise times were shorter in both unipodal stances with respect to their bipodal homologues. An increase in COP path length, velocity and sample entropy was observed when the support area was reduced. All parameters showed good reliability in both directions at all stances. This framework could be used in the clinical practice to assess dynamic postural control capabilities in patients whose balance is pathologically affected.

## 1. Introduction

Postural control is the set of mechanisms that provide stability to the human body and allow it to adopt and maintain different postures. It is a complex skill that is required for everyday activities that uses multiple resources, such as representations of biomechanical limits, dynamic control, orientation in space, sensory integration, movement strategies and associated cognitive processes [1]. These resources can be affected by lower limb or sensory system problems, by diseases like diabetes, and by neuromuscular pathologies. The effects of the lack of resources associated with postural control has been analysed in several studies [2], [3], [4]. The influence of diseases such as Parkinson’s [5], [6], [7], [8] and Alzheimer’s [9], or simply aging on postural control [10], [11] has been investigated as well. Furthermore, many studies characterised static postural control in bipodal and unipodal postures [12], [13], [14], [15], [16] and the response of postural control to external perturbations [17].

Beyond that, very little has been investigated on the response of dynamic postural control to voluntary and stable weight shifting stances. Dingenen et al. [18] modified the time to stabilization method to analyse postural stability during the transition task from double-leg stance to unipodal stance with chronic ankle instability volunteers. Additionally, De la Torre et al. [19] proposed a rhythmic weight shift test to analyse dynamic changes while volunteers followed linear trajectories. In [20], a novel conceptual framework for studying human postural control was presented, using the center of pressure (COP) velocity autocorrelation function, analyzing its variations under alterations in the visual and stance conditions, and comparing its results to Stabilogram Diffusion Analysis (SDA), considering in their study alterations in the same position.

An alternative approach has been carried out by Picoli et al. [21], [22], who analyzed the distribution of the COP reference crossing events in uni- and bi-podal stances. In [23], the authors introduced the analysis of the variations in the coefficients from a fractional Brownian motion, in order to model the postural corrections in unipodal or bipodal stances. A similar approach has been presented in [21] and [22], where the focus was spotted over a COP reference zero crossing time variance and distribution, for bipodal and unipodal stances. Beyond that, very little has been investigated on the response of dynamic postural control to voluntary and stable postural changes. In particular, in [24]. The authors analyzed the incidence of the initial bipodal stance width to the unipodal balance time. In such a work, authors analyzed the relation between these variables, but they did not characterise the involved dynamics.

In this context, the main objective of this study was the characterization of the dynamic postural control during weight load shifting with and without support surface reduction, with temporal metrics commonly used in linear control systems identification. For this purpose, an experimental protocol was devised to record the COP coordinates during the execution of dynamic postural control tasks in humans. Global, structural, and temporal variables were calculated from the COP coordinates. Additionally, the validity and reliability of the assessed variables parameters were determined.

## 2. Materials and Methods

### 2.1 Participants

Fourteen healthy volunteers between 18 and 30 years old participated in the experiment. None of them reported recent injuries to their lower limbs or neurological diseases affecting postural control. The protocol was evaluated and approved by the Central Committee of Bioethics in Biomedical Practice and Research of the province of Entre Ríos. All participants provided written informed consent prior to participation, and the Declaration of Helsinki was respected.

### 2.2 Experimental design

Two experimental sessions were carried out, separated by one week. The tasks performed by the volunteers were: bipodal (B) stance with asymmetrical unloading of the body weight in medial-lateral direction (BR with displacement to the right, BL with displacement to the left), bipodal on tiptoes (T) and unipodal (U) with each of the lower limbs supported (UR for right limb support, UL for left limb support). All tasks began with the volunteer adopting a bipodal stance with a comfortable foot separation. For this experiment, 30 trials of each task were performed per session, divided into two blocks of 15 trials each, with a rest time of 5 s between trials and 2 to 3 min between blocks, as shown in Fig. 1. The order of the tasks was randomized per block.

**Fig. 1.**
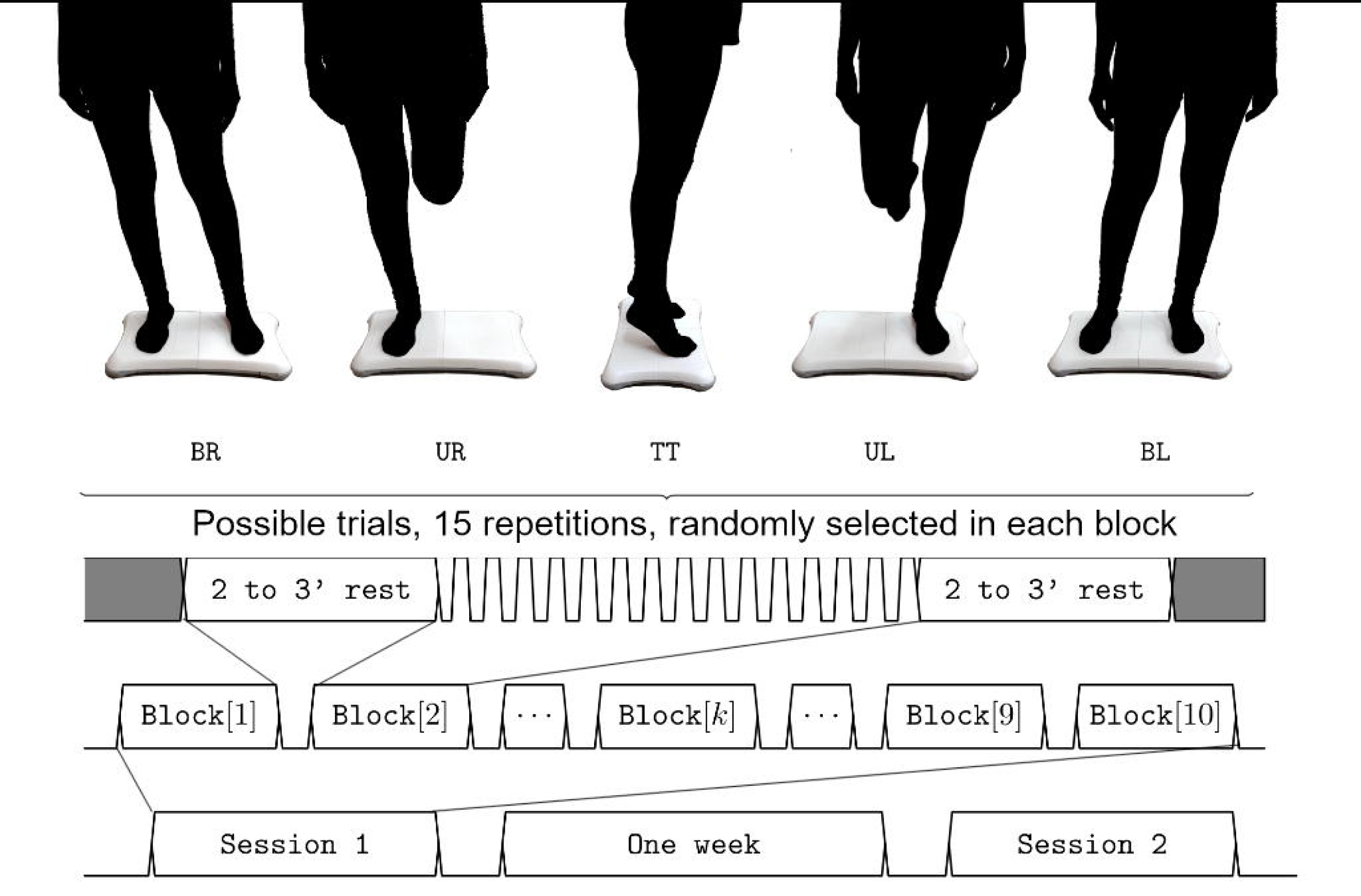
Temporal diagram of the experimental protocol.

A software interface was generated to obtain synchronised recordings of biomechanical signals from a WBB. This interface made it possible, through its configuration, to comply with the guidelines established in the experimental protocols. These protocols contemplated the performance, by healthy volunteers, of tasks such as bipodal posture (BR and BL) and postures with decrease of support surface (UR, UL and T). The recording of the COP coordinates was performed in two sessions separated by a period of one week, which allowed the evaluation of the reliability of the analysed biomechanical parameters.

### 2.3 Data recording

A Wii Balance Board (WBB) was used for balance assessment [25], [26], [27], [28], [29]. A software interface was implemented to get the signals of the four sensors of the platform through Bluetooth® protocol and store them in text files. The sampling frequency was 64 Hz. The interface also had configuration options related to the protocol such as: number of trials, trial duration, tasks type and feedback modality (auditory or visual).

### 2.4 Data pre-processing

A fourth order Butterworth low-pass filter with a cut-off frequency of 12 Hz was used to remove high-frequency noise from the platform signals obtained by the interface [26]. After that, the equations described in [25] and in [28] were applied to these signals to calculate the COP.

Trials were divided into two segments: the first 2 s, corresponding to the transient response between an initial state (i.e. bipodal stance) and the final state represented by the postures BR, BL, UR, UL and T; and the remaining 3 s that comprise the response of the subjects in steady state in the new posture or final state. A series of temporal variables that characterize the dynamics of the COP were calculated with the samples corresponding to the first segment, and global and structural variables were calculated with data from the second segment.

### 2.5 Temporal variables

Delay time (T50) is the time required for a system’s response to reach 50% of its final value, while rise time (T10-90) is the time required for the response to move from 10% to 90% of its final value [30]. These parameters are frequently used to describe first and second order linear systems, commonly described in the literature. Delay time and rise time were calculated as temporal variables to characterize the transient response of the postural control system on both AP and ML directions.

### 2.6 Global variables

Mean value of the COP position, standard deviation and maximum excursion, on anterior-posterior and medial-lateral directions, COP path length (P) and COP velocity were calculated as proposed by [31].

### 2.7 Structural variable

Sampling entropy (SE) was considered as a structural variable, taking into consideration that entropy measures have been previously used for the analysis of COP variations under different postural control situations [32]. SE was obtained from the negative natural logarithm of the conditional probability of a data sequence. High SE values come from sequences with low probability of repetition, thus representing low regularity and high complexity in the data [33].

### 2.8 Statistics

Linear mixed models determined differences in temporal, global and structural variables, in which *posture* (levels: B, U and T) and *direction* (levels: R and L) were considered as fixed factors (whenever appropriate). Main effects and interactions were analysed, and random intercepts were allowed for subjects. P-values less than 0.05 were considered statistically significant. Tukey’s test was used for post-hoc comparisons.

Bland-Altman analysis was performed to assess the reliability of the outcome variables. This analysis is based on the evaluation of the mean against the difference of two repeated measurements on the same subject. The limits of agreement were also derived as the mean difference (bias) ± 1.96 times the standard deviation of the differences between measurements. The limits of agreement delimit the range within which 95% of the differences between measurements lie under normal conditions, which can be interpreted as the maximum expected difference between measurements due to measurement error, in the absence of other factors [34].

## 3. Results

### 3.1 Temporal variables

#### 3.1.1 Delay time

T50 mean values of the fourteen volunteers and the corresponding boxplot for each session and posture in ML and AP directions are shown in Fig. 2. Posture had a significant effect on the T50 in the ML direction (F_1,94_=70.72, p<0.001). Post hoc tests showed shorter T50 times in U compared to B or T (mean difference: 0.22 ± 0.02 s, t_94_=8.41, p<0.001).

**Fig. 2:**
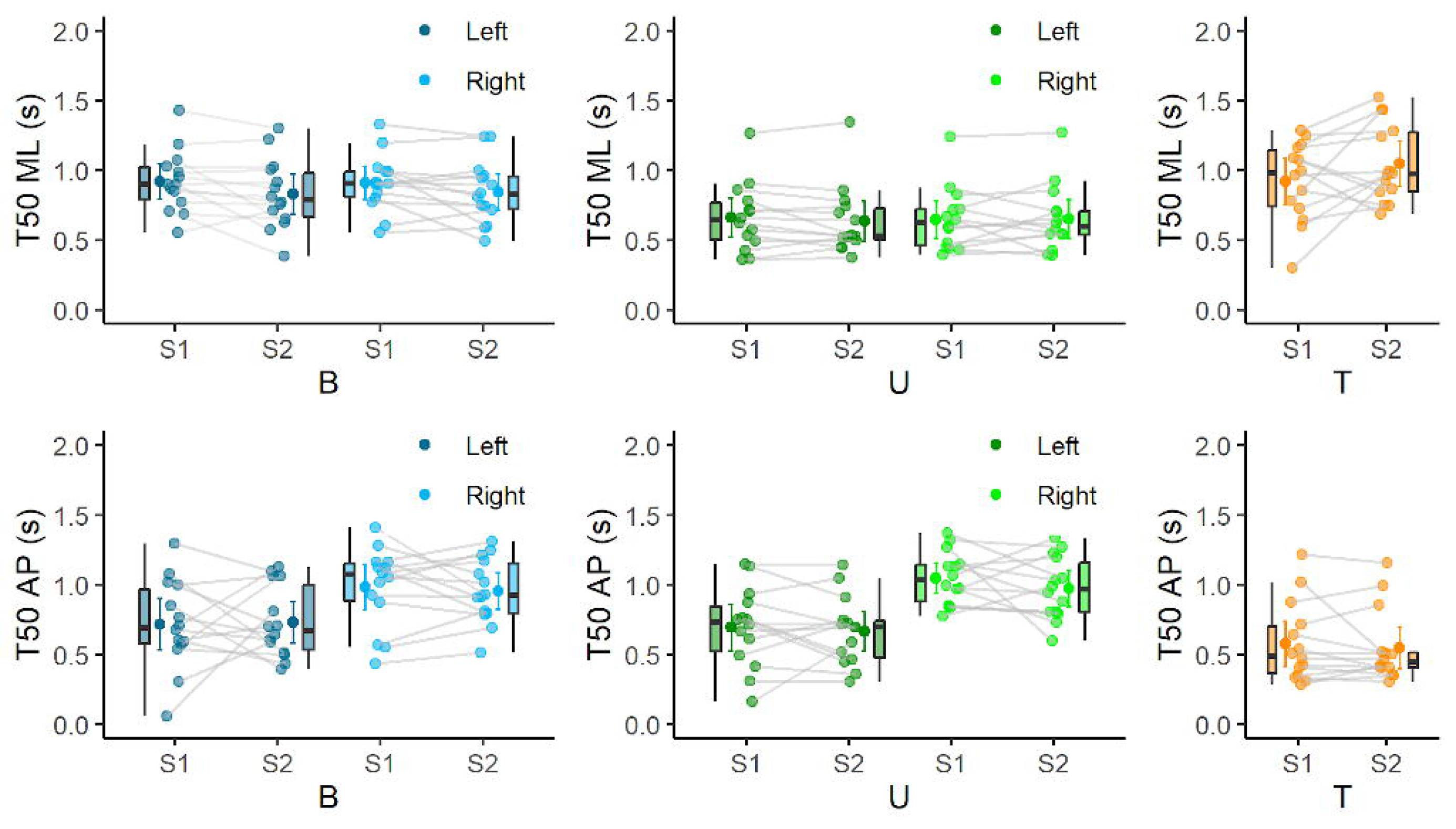
Each dot represents each average T50 for each session in ML and AP directions. Boxes with the 50% of the average measures for each individual. T50 mean values of the fourteen volunteers and the corresponding boxplot for each session (S1 and S2) and posture (B, U and T) supported with the right, left or both lower limbs in ML (above row) and AP (below row) directions.

A particular behaviour was found in the side analysis, with a significant effect on the T50 in AP direction (F_1,94_=41.94, p<0.001). Post hoc tests showed a greater mean value of the T50 in AP direction comparing the same task executed with different limb, BR against BL and UR against UL postures (mean difference: 0.28 ± 0.04s, t_94_=6.48, p<0.001).

Regarding T posture, delay time mean value in ML direction was 0.98s and 0.56s in AP direction.

No significant difference between sessions was observed for any posture or direction. An increase in the limits of agreement in the direction of least displacement (AP for B and U, and ML for T), from 0.25 s to approximately 0.5 s, was observed.

#### 3.1.2 Rise time

Posture had a significant effect on the T10-90 in the ML direction (F_1,94_=59.62, p<0.001). Post hoc tests showed shorter T10-90 times in U compared to B or T (mean difference: 0.23 ± 0.03 s, t_94_=7.72, p<0.001).

In B postures, similar mean values of T10-90 were observed in both directions, whereas, higher mean values were displayed by the postures with decreased support surface (U and T) in the direction of less displacement (F_1,95_=25.19,p<0.001), as shown in Fig. 3.

**Fig. 3:**
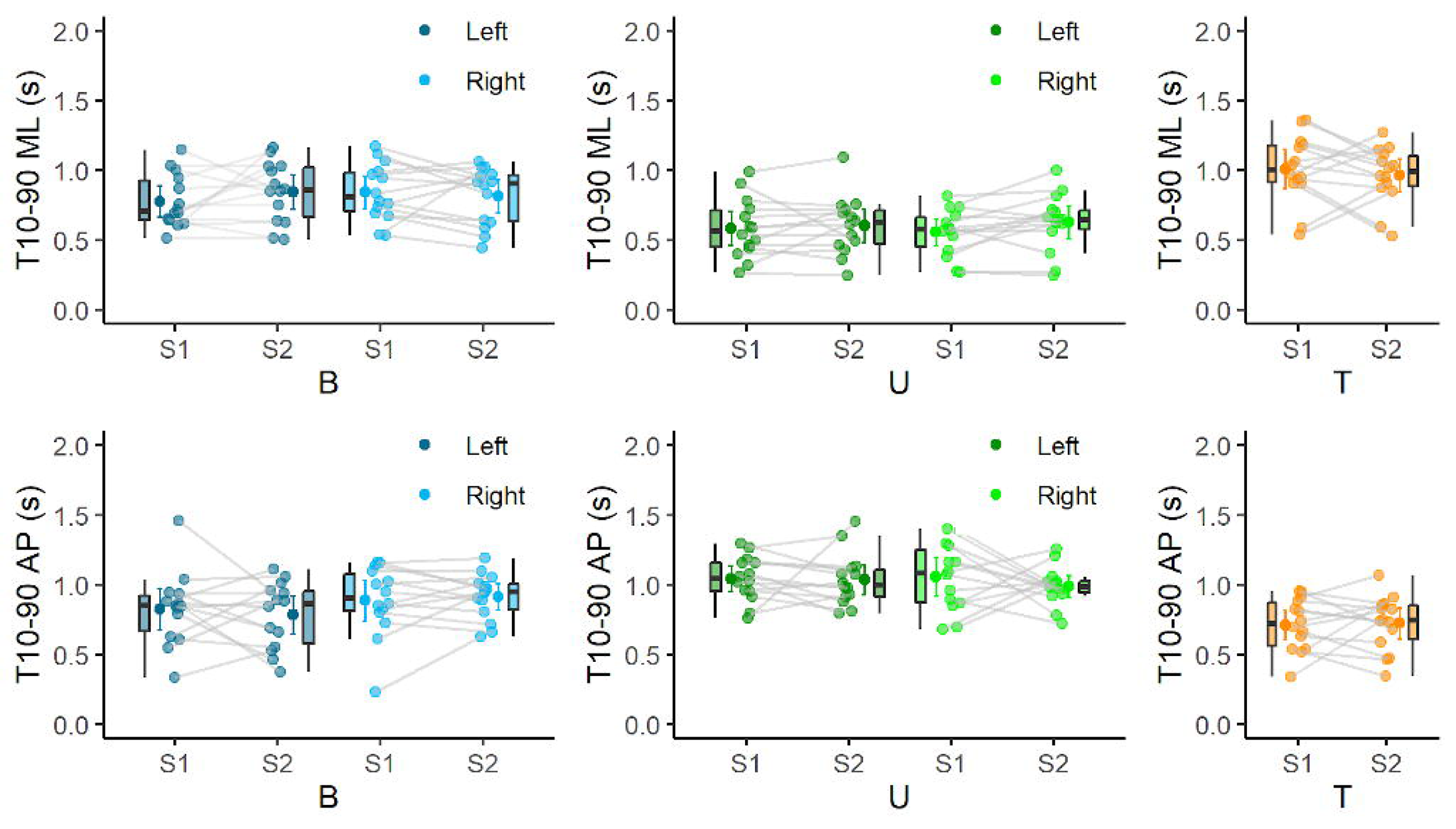
Each dot represents each average T10-90 for each session in ML and AP directions. Boxes with the 50% of the average measures for each individual. T10-90 mean values of the fourteen volunteers and the corresponding boxplot for each session (S1 and S2) and posture (B, U and T) supported with the right, left or both lower limbs in ML (above row) and AP (below row) directions.

From the Bland Altman analysis, it can be seen that the bias was close to zero and the limits of agreement were within approximately 0.5s for all stances and directions.

#### 3.1.3 Global Variables

Mean values of COP in ML direction showed no differences. In AP direction, an increase in mean value in U postures was observed with respect to B postures ( *F*_1,94_ = 27. 23 *p* < 0. 001). A similar behavior was observed in the standard deviation in the same analysis (*F*_1,95_ = 101. 97, *p* < 0. 001). Moreover, lower values of maximum displacement were noted in B postures with respect to U postures on both directions (*F*_1,95_ = 81. 91, *p* < 0. 001 ML; *F*_1,95_ = 136. 74, *p* < 0. 001 AP).

As is exhibited in Fig. 4, U postures generated higher path length and velocity values than B postures ( *F*_1,95_ = 154. 45, *p* < 0. 001).

**Fig. 4:**
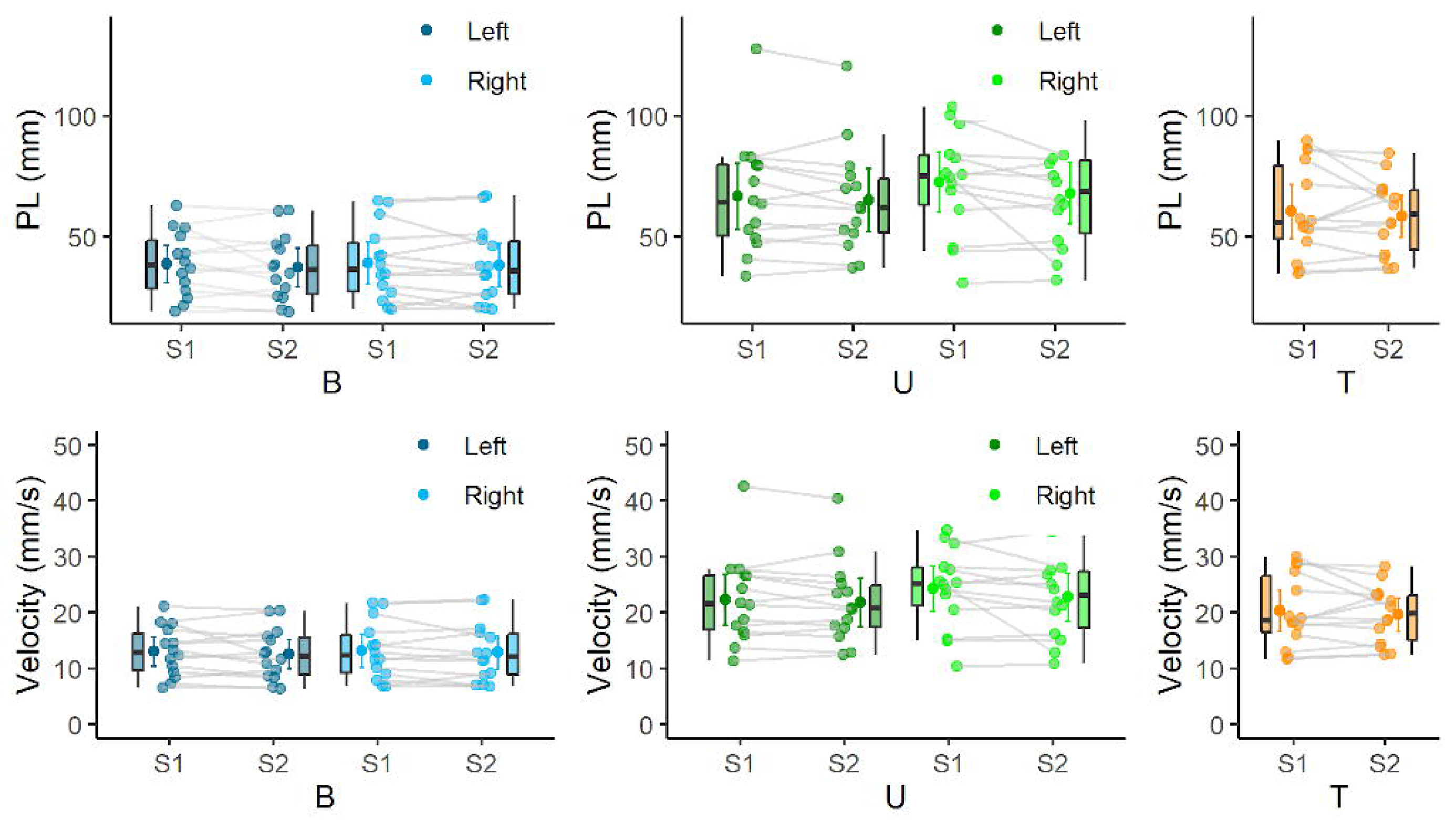
PL (above row) and velocity (below row) mean values of the fourteen volunteers and the corresponding boxplot for each session (S1 and S2) and posture (B, U and T) supported with the right, left or both lower limbs.

The bias values obtained from the Bland-Altman analysis were not relevant for all the global variables and the limits of agreement were lower, a few millimetres, than the mean values of the corresponding variable.

### 3.2 Structural variables

The mean values of SE for B postures were the lowest from all considered postures (*F*_1,95_ = 214. 41, *p* < 0. 001 for SE ML y *F*_1,95_ = 92. 29, *p* < 0. 001 for SE AP), as shown in Fig. 5. The mentioned difference in mean values is enlarged comparing ML direction with AP direction. These findings indicate an increase in complexity of the temporal series when the support surface is reduced.

**Fig. 5:**
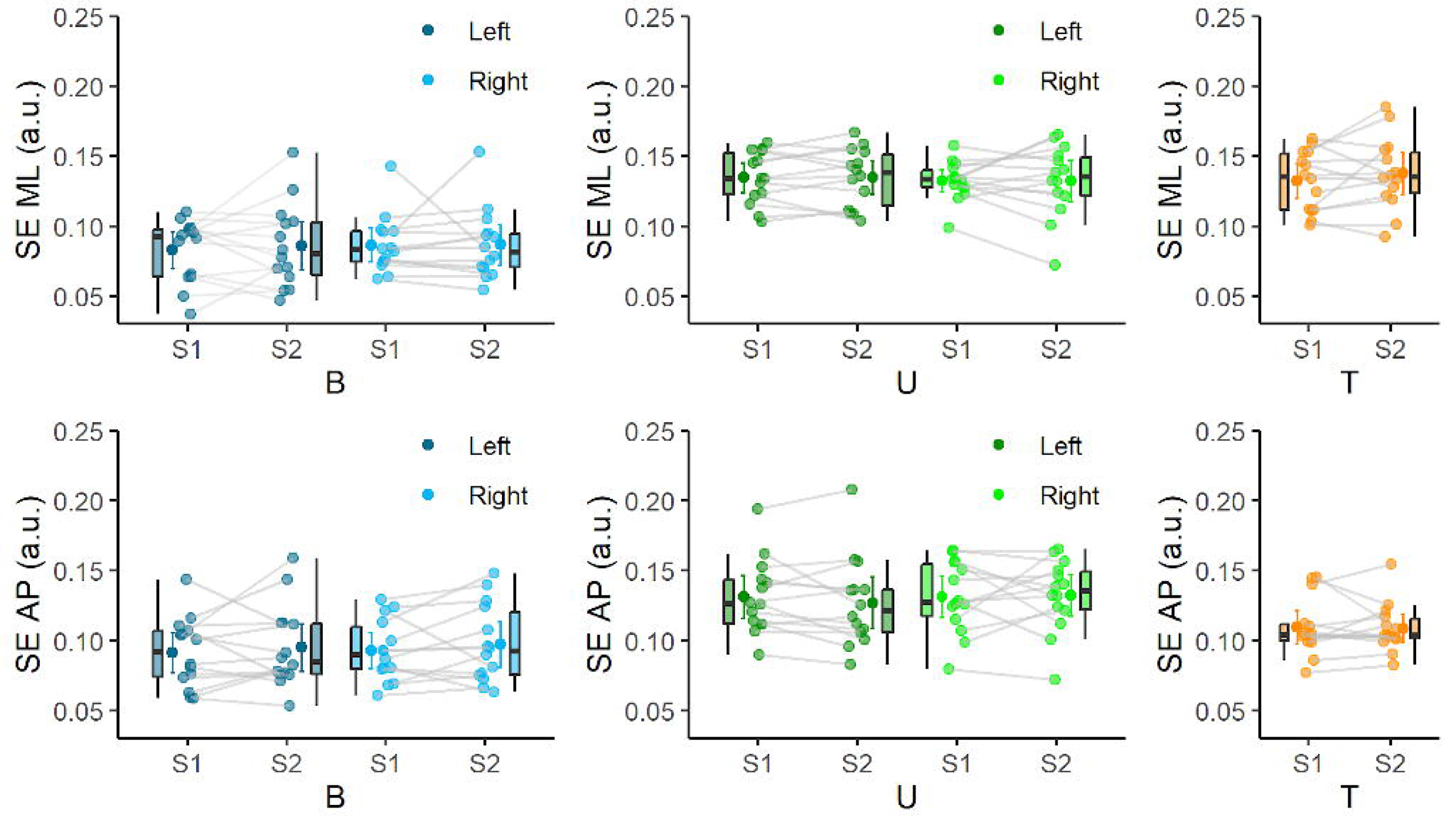
SE mean values of the fourteen volunteers and the corresponding boxplot for each session (S1 and S2) and posture (B, U and T) supported with the right, left or both lower limbs in ML (above row) and AP (below row) directions.

The Bland-Altman analysis of the SE values manifested no bias between sessions and the limits of agreement were close to 0.04 value.

## 4. Discussion

### 4.1 Temporal variables

Different parameters calculated when adopting a variety of postures (unipodal or bipodal) were previously described in detail under the influence of diverse factors [35]. Richardson et al. [24] analysed the incidence of the initial bipodal stance width to the unipodal balance time. The authors analysed the relationship between these variables, but they did not characterise their dynamics. Additionally, de la Torre et al. [19] proposed a rhythmic weight shifting test to analyse dynamic changes while volunteers followed linear trajectories. Furthermore, Dingenen et al. [18] measured the duration of the contralateral push-off movement and the peak COP velocity to analyse postural stability during the transition task from double-leg stance to single-leg stance with chronic ankle instability volunteers. However, very little is reported on the transitions between postures, limiting the dynamics analysis only to the steady state, neglecting the transient information. In the present study new parameters in this research field were used to characterise weight shifting stances, namely, delay and rise time.

Results showed a significant difference in T50 (between 23 and 33 % on average) in the AP direction when the same task was performed with the dominant limb or with the non-dominant limb. A similar effect was observed for the rise time, but with a smaller effect size. This asymmetry in neuromuscular control between dominant and non-dominant side has been previously established in Promsri [36]. However, this is not fully consistent with the results in Wieslaw Błaszczyk et al. [37], where no significant differences were found in a similar protocol. These differences could be associated with a number of factors, such as the sample composition, or differences in the specific motion patterns of the volunteers.

Our results showed a decrease in the T50 and T10-90 in both unipodal stances with respect to their bipodal homologues. In the ML direction, resulting in a faster response when a weight shifting with reduction in the support area was performed. This is consistent with the results in Błaszczyk et al. [37], since the neuromuscular control mechanisms involved in the bipodal and unipodal stances are characterised by different dynamics and kinematic profiles. The T50 and T10-90 reductions in these more complex tasks can be associated with the increase in the volunteer conscientiousness, in a similar way as reported in [41] for different stance conditions. Finally, temporal parameters were shown to be reliable in both directions and all stances.

### 4.2 Global variables

Hernandez et al. [20] presented a novel conceptual framework for studying human postural control, using the centre of pressure (COP) velocity autocorrelation function (COP-VAF), analysing its variations under alterations in the visual and stance conditions, and comparing its results to the Stabilogram Diffusion Analysis (SDA). An alternative approach has been proposed by Picoli et al. [21], who analysed the distribution of the COP reference crossing events in uni- and bi-podal stances. In time, Burdet and Rougier [23] introduced the analysis of the variations in the coefficients from a fractional Brownian motion, in order to model the postural corrections in unipodal or bipodal stances. In these studies, a significant difference on the analysed parameters between the unipodal stance and the bipodal stance was reported.

In this study, the decrease of the support surface (UR and UL stances) generated a similar instability in both directions (ML and AP) and the conservation of the support surface (BR and BL stances) generated a greater deviation in the ML direction. An increase in COP path length and velocity was observed when the task difficulty was increased by a decrease in the support surface, in accordance with previous studies [38].

The Bland-Altman analysis of the global variables showed in general acceptable reliability. A small bias was detected in the mean values of the T stance in the AP direction, which would indicate that the values of the first session were, on average, higher than those of the second session. This could be explained by the fact that this is a complex stance in which the subjects may have modified the displacement in the AP direction mainly because they did not have any predefined distance to reach in this direction in the experiment or that in the second session they made a more controlled change of stance with respect to the first session.

### 4.3 Structural variables

Ahmadi et al. [39] studied the discriminatory ability of sample entropy to compare treadmill walk only with a dual-task condition. Regarding human postural sway during quiet standing, Ramdani et al. [40] computed sample entropy to successfully discriminate two sensory conditions (eyes-open and eyes-closed) in a group of healthy young adults. Comparably with the reported studies, the values obtained for the structural variable would indicate an increase in the complexity of the COP time series when a lower support surface position is performed, a situation that occurs in the ML and AP directions. This variable proved to be reliable and limits of agreement were established above 0.04 for series of 3s duration.

### 4.4 Limitations

The experimental protocol allowed us to explore the application of typical dynamic systems concepts to postural transition analysis, although the sample size and composition is not sufficient to generalise to a broad population. While there exists literature that evaluates the COP during a reduction in the support area, there is no standardised protocol for the transition from a bipodal to unipodal stance. Indeed, the absence of a normalised motion pattern for these tasks does not enable a direct comparison between our results and those already in the literature.

## 5 Conclusions and future directions

Tasks involving weight load shifting with and without support surface reduction were performed faster with increasing levels of complexity. This can be explained considering that in these activities, more conscious neural circuits should be activated, since it might be associated with more falling risk. The utilisation of temporal parameters in the clinical practice might introduce new insights in how injured patients are evaluated and how their rehabilitation evolves. In order to proceed in such a direction, wider studies should be performed, considering the target population of the potential application.

## Author Contributions

Esteban OSELLA: Visualization, Writing - Original Draft, Resources, Methodology, Formal analysis, Review & Editing. Ricardo Alfredo RETTORE: Data Curation, Resources, Investigation, Software. Paola CATALFAMO: Supervision, Writing - Review & Editing, Conceptualization, Methodology. José Alberto BIURRUN MANRESA: Conceptualization, Methodology, Formal analysis, Supervision, Writing - Review & Editing. Yanina Verónica ATUM: Project administration, Funding acquisition, Visualization, Writing - Original Draft, Resources, Investigation, Formal analysis, Software, Conceptualization, Methodology.

**Esteban OSELLA** received the biomedical engineering degree, and the PhD degree in engineering. He is at Institute of bioengineering and Bioinformatics (IBB - CONICET-UNER) and FI UNER. His research interests include control systems applied to lower limb rehabilitation strategies.

**Ricardo Alfredo RETTORE** received the biomedical engineering degree. He is at Institute of bioengineering and Bioinformatics (IBB - CONICET-UNER) and FI UNER. His research interests include control systems applied to lower limb rehabilitation strategies.

**Paola CATALFAMO** received the biomedical engineering degree, and the PhD degree in engineering. He is at Institute of bioengineering and Bioinformatics (IBB - CONICET-UNER) and FI UNER. Her research interests include prosthetics gait analysis and biomechanics.

**José Alberto BIURRUN MANRESA** received the biomedical engineering degree, and the PhD degree in engineering. He is at Institute of bioengineering and Bioinformatics (IBB - CONICET-UNER) and FI UNER. His research interests include human neurophysiology, with particular emphasis on the study of nociception and pain.

**Yanina Verónica ATUM** received the biomedical engineering degree, and the PhD degree in engineering. He is at Institute of bioengineering and Bioinformatics (IBB - CONICET-UNER) and FI UNER. Her research interests include neurosciences, mainly in the area of biosignal processing.

## Data Availability

All data produced in the present study are available upon reasonable request to the authors

---

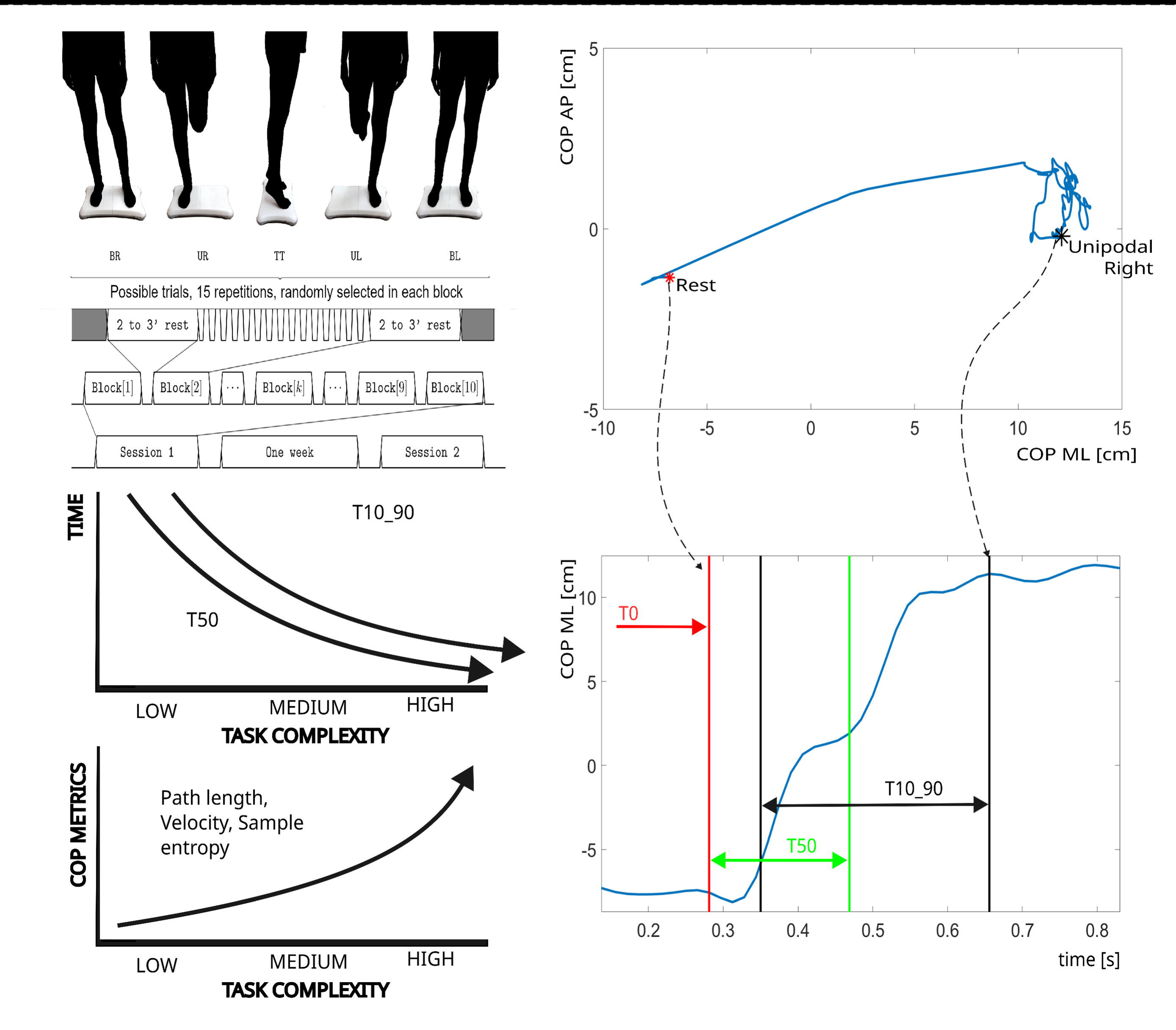

